# Natural killer cells are a potential source of oxidative stress in inclusion body myositis

**DOI:** 10.1101/2025.05.05.25326736

**Authors:** Lyn Eigenfeldt, Marc Pawlitzki, Sven G. Meuth, Tobias Ruck, Christopher Nelke

## Abstract

Inclusion body myositis (IBM) is the most common idiopathic inflammatory myopathy in individuals over 50 years of age. IBM is considered a T cell-mediated disease. In contrast, the role of natural killer (NK) cells remains understudied. Here, we characterized the phenotype and function of peripheral NK cells in 22 IBM patients and 22 age-matched non-diseased controls. Flow cytometry revealed stable overall NK cell frequencies but a shift toward a more cytotoxic phenotype in IBM, with elevated NKG2D expression and increased granzyme A and B levels in CD56^dim^ NK cells. Functional assays demonstrated that NK cells from IBM patients exhibited enhanced cytotoxicity against primary human muscle cells. These findings suggest that NK cells may contribute to muscle fiber injury in IBM through NKG2D-driven cytotoxicity and reactive oxygen species generation. Our study highlights NK cells as potential amplifiers of inflammation and oxidative stress in IBM.

## Introduction

Inclusion body myositis (IBM) is the most common idiopathic inflammatory myopathy (IIM) in individuals over 50 years of age ^1^. Clinically, IBM presents with progressive muscle weakness, particularly affecting the quadriceps and finger flexors, leading to substantial disability. Despite its prevalence, effective therapeutic options remain limited ^2^.

The pathogenesis of IBM is complex, involving both inflammatory and degenerative processes. A hallmark feature is the invasion of muscle fibers by cytotoxic CD8+ T cells, which are clonally expanded and directed against unknown muscle antigens ^2^. These T cells exhibit features of chronic activation and contribute to muscle fiber damage.

While T cells have been extensively studied in IBM, the contribution of natural killer (NK) cells remains less clear. NK cells are cytotoxic lymphocytes capable of releasing granzyme and perforin, as well as producing pro-inflammatory cytokines and ROS ^3^. In chronic inflammatory settings, they can adopt a tissue-resident phenotype and contribute to sustained immune activation. Given their ability to generate oxidative stress and modulate T cell responses, NK cells represent a plausible contributor to the inflammatory niche observed in IBM. However, recent studies have reported that broader peripheral NK cell subsets are unchanged in IBM patients ^4^.

In this study, we aim to further characterize the phenotype and function of NK cells in IBM, exploring their potential contribution to muscle inflammation and oxidative stress. We position that NK cells may augment T cell–mediated cytotoxicity and perpetuate ROS-driven tissue damage in IBM.

## Material and Methods

### Study cohort

We included 22 patients with confirmed diagnosis of IBM according to national guidelines and 22 non-diseased, age-matched controls (NDCs). Detailed patient characteristics are given in **Table 1**. Disease duration of IBM patients was defined as the time in months between symptom onset and blood sampling. Manual muscle testing of 8 muscle groups (MMT-8) was used for clinical assessment. The antibody status was determined using an EUROLINE Blot assay (Euroimmun). Creatine kinase (CK) levels were measured during routine laboratory testing at the same time as the performance of study blood sampling. The local ethics committee (2016-053-f-S) approved blood sampling, and all subjects provided written informed consent before entering the study. This trial was conducted in accordance with the Declaration of Helsinki.

**Table 1:** Demographics and baseline disease characteristics. Baseline is defined as the time of blood draw. NDC = non-diseased control, IBM = inclusion body myositis.

|  | IBM | NDC |
| --- | --- | --- |
| Total number | 22 | 22 |
| Median age diagnosis (years, range) | 64 (39 – 84) | 62 (35 – 87) |
| Median disease duration (years, range) | 1.7 (0.2 – 6.4) | - |
| Female (n, %) | 10 (45%) | 12 (54%) |
| Antibody status (n, %) | Anti-cN1A 8 (36%) | - |
| Treatment naïve (n, %) | 16 (72%) | - |
| Intravenous immunoglobulins (n, %) | 6 (27%) | - |

### Sampling and flow cytometric analysis of peripheral blood mononuclear cells

Whole blood samples were collected from study subjects. Peripheral blood mononuclear cells (PBMC) were isolated by Ficoll (Sigma-Aldrich) density gradient centrifugation. Samples were stored in liquid nitrogen as previously described ^5^. Thereafter, PBMCs were centrifuged at 300×g for 5 minutes and suspended in phosphate buffered saline (PBS, Sigma-Aldrich) supplemented with 2% heat-inactivated fetal bovine serum (FBS, GE Healthcare) and 2 mM ethylenediaminetetraacetic acid (EDTA, Sigma-Aldrich) and incubated with fluorochrome-conjugated antibodies at 4°C for 30 minutes. Staining of chemokine receptors was performed at 4°C for 30 minutes. PBMCs were washed and additionally stained with the Zombie NIR or Aqua Fixable Viability Kit (Biolegend) according to the manufacturer’s manual to distinguish between dead and living cells. Only samples with a viability of >85% were permitted. PBMCs were washed and suspended in PBS/FBS/EDTA and analyzed by flow cytometry (FC) using a CytoFlex Flow Cytometer (Beckman Coulter). For staining of intracellular proteins (perforin and granzyme, Gr) components from the BD Cytofix/Cytoperm™ Kit (BD Biosciences) were used according to the manufacturer’s manual. Data were analyzed using Kaluza Flow Cytometry Analysis software version 2.1 (Beckman Coulter).

### Gating strategy

Immune cell subsets in the peripheral blood were identified using the following markers: Lymphocytes: FSC vs. SSC, B cells: CD19^+^ CD3^-^ Lymphocytes, T cells: CD3^+^ CD56^-^ Lymphocytes, CD4: CD4^+^ CD8^-^ T cells, CD8: CD8^+^ CD4^-^ T cells, NK cells: CD56^+^ CD3^-^ Lymphocytes, CD56^dim^: CD56^dim^ CD16^+^ NK cells, CD56^bright^: CD56^bright^ CD16^-^ NK cells.

### NK-cell mediated cytotoxicity assessment

Primary human muscle cells (PHMCs) were derived from healthy donor biopsies and expanded in growth medium until 70–80% confluence, as previously described ^6^. NK cells were isolated from PBMCs obtained from either IBM patients or NDCs, as described above. Isolation was performed using flow cytometry, gating on CD3^-^CD56^+^ cells to exclude T cells and enrich for the NK cell population. Purity of sorted cells was confirmed to be >95%. For co-culture experiments, PHMCs were seeded in 24-well plates and co-incubated with NK cells at varying effector-to-target (E:T) ratios as indicated. Co-cultures were maintained in cell culture medium for 24 hours. After 24 hours of incubation, cytotoxicity was assessed using the Invitrogen™ LIVE/DEAD™ Cell-Mediated Cytotoxicity Kit for mammalian cells. Fluorescence-based detection of dead target cells was performed using using microscopy. 50 cells per sample were chosen at random and the percentage of live or dead cells were counted. All experiments were performed in at least three biological replicates.

### Statistical analysis

Statistical Analysis was performed using GraphPad Prism 10.1 (GraphPad Software, Inc., San Diego, CA). Data are presented as median (IQR = interquartile range), mean (standard deviation = SD) or n (%). Differences between groups were analyzed using unpaired Student’s t test or Mann–Whitney-U test as appropriate. Differences between groups and a hypothetical mean were analyzed using a one-sample t test. Differences were considered statistically significant with the following P-values: *p < 0.05, **p < 0.01, ***p < 0.001 and ****p < 0.0001.

### Data availability

All data associated with this study are present in the paper. The data assessed in this study are available from the corresponding author on reasonable request.

## Results

### NK cells are cytotoxic in IBM

To investigate the role of peripheral NK cells, we recruited a cohort of IBM patients fulfilling the ENMC histopathological criteria ^7^ alongside age-matched non-diseased controls (NDCs) with no known medical conditions (**Table 1**). Age-matching was implemented to control for age-related immune changes (**Fig. 1A**). Clinical metadata, including disease duration at the time of study inclusion, were recorded (**Fig. 1B**).

**Figure 1:**
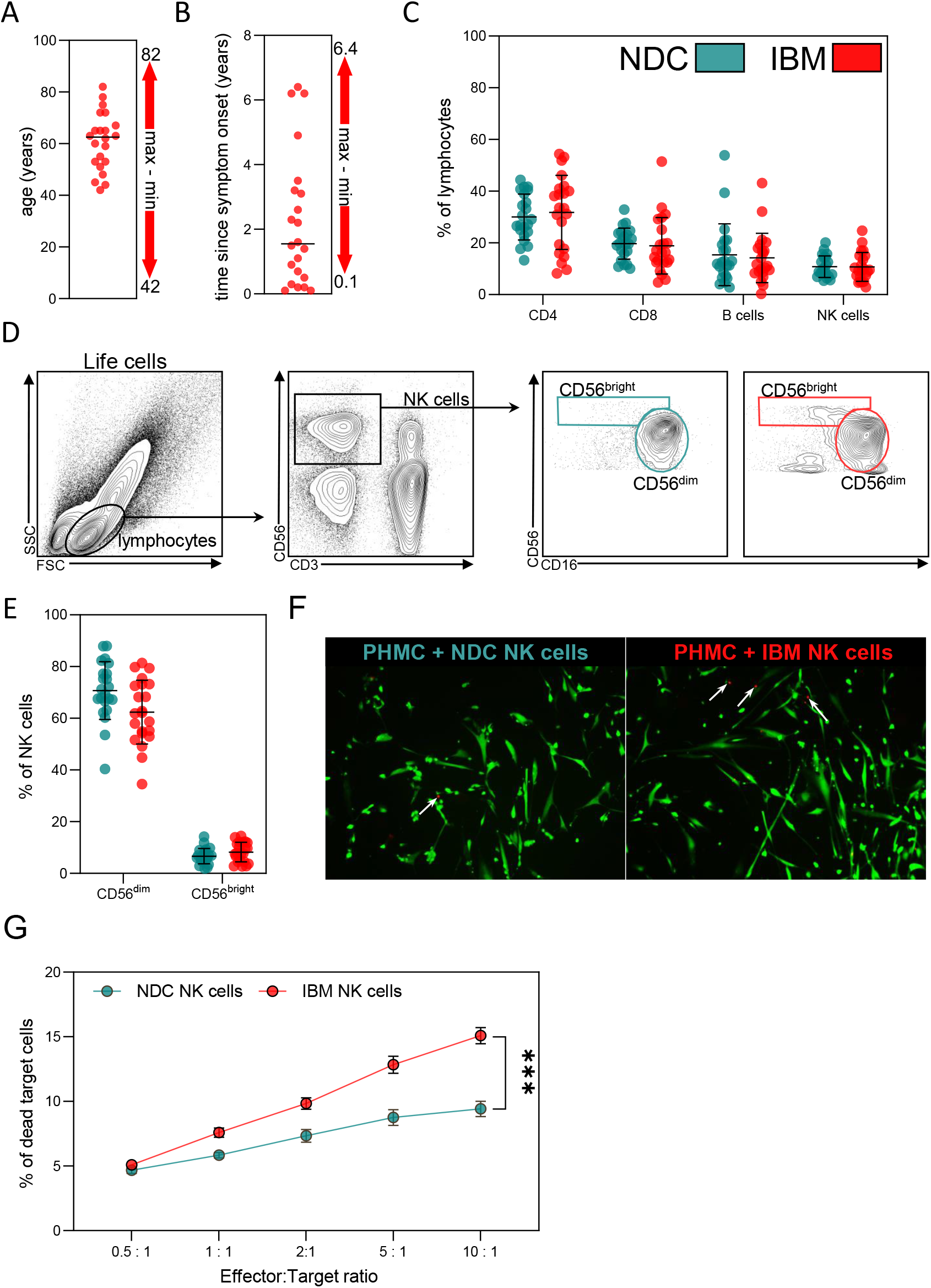
NK cells are cytotoxic in IBM. (**A**) Dot plot indicated the age at study inclusion for the IBM patients. (**B**) Dot plot indicated the time since symptom onset in years at study inclusion. The study inclusion is the time of blood draw for the assessment of NK cell populations. (**C, D**) Cells from peripheral blood were analyzed by flow cytometry. Total leukocytes were identified by forward scatter channel (FSC) characteristics and as CD45 expressing cells. From these, lymphocytes were selected based on CD56 signal and CD14 expression. Within lymphocytes, B cells were identified as CD19^+^CD138^-^ cells. Lymphocytes were divided into CD3^+^CD56^+^ NKT cells, CD3^+^CD56^-^ T cells, and CD56^+^CD3^-^ NK cells. NK cells were further separated into CD56^dim^CD16^high^ and CD56^bright^CD16^dim^/^-^ NK-cell subsets. T cells were split into CD4^+^CD8^-^ and CD8^+^CD4^-^ T cells. (**E**) Relative cell numbers of NK cell subsets. (**F**) Exemplary live/dead-staining for PHCMs incubated with varying numbers of NK cells. Dead cells are indicated as red dots. Live cells are green. (**G**) Percentage of dead target cells (PHMCs) at each Effector (E, NK cell) to Target (T, PHMC) ratio. Differences were determined by the two-sided student’s T-test. Abbreviations: FSC = forward scatter channel, IBM = inclusion body myositis, NDC = non-diseased control, NK = natural killer, PHMC = primary human muscle cell, *** p < 0.001, * p < 0.05

We first performed multicolor flow cytometry to characterize immune cell subsets in peripheral blood. Frequencies of adaptive immune cells – including CD4^+^ T cells, CD8^+^ T cells, and B cells – did not differ significantly between IBM and NDCs (**Fig. 1C**). Total NK cell frequencies were also comparable across groups. However, phenotypic analysis of NK cell subsets revealed a trend toward redistribution: the proportion of CD56^dim^ NK cells was reduced in IBM, with a relative increase in CD56^bright^ NK cells (**Fig. 1D, E**). Although this shift did not reach statistical significance due to inter-individual variability, it is consistent with reports from other autoimmune conditions where peripheral CD56^dim^ NK cells are depleted ^8,9^. These findings support the notion that overall NK cell numbers remain stable in IBM, but their composition may be altered.

We next examined whether NK cell cytotoxicity is functionally altered in IBM. To this end, we isolated NK cells from the peripheral blood and co-cultured them with primary human muscle cells (PHMCs) at varying effector-to-target (E:T) ratios. Cytotoxicity was assessed after 24 hours using a fluorescence-based viability assay (**Fig. 1F, G**). Strikingly, NK cells derived from IBM patients consistently exhibited higher cytotoxic activity compared to those from NDCs, as indicated by increased numbers of dead target cells. This suggests that although peripheral NK cell numbers are unchanged, their functional state in IBM is skewed toward enhanced cytotoxicity.

### NK cell phenotypes in IBM

CD56^dim^ NK cells are the primary cytotoxic subset and have been shown to accumulate at sites of inflammation. To explore their potential role in IBM, we analyzed the expression of surface receptors that shape NK cell activation and effector function. We focused on the CD3^-^CD56^+^ NK cell population (**Fig. 2A**) and profiled cell surface markers in the CD56^dim^ subset.

**Figure 2:**
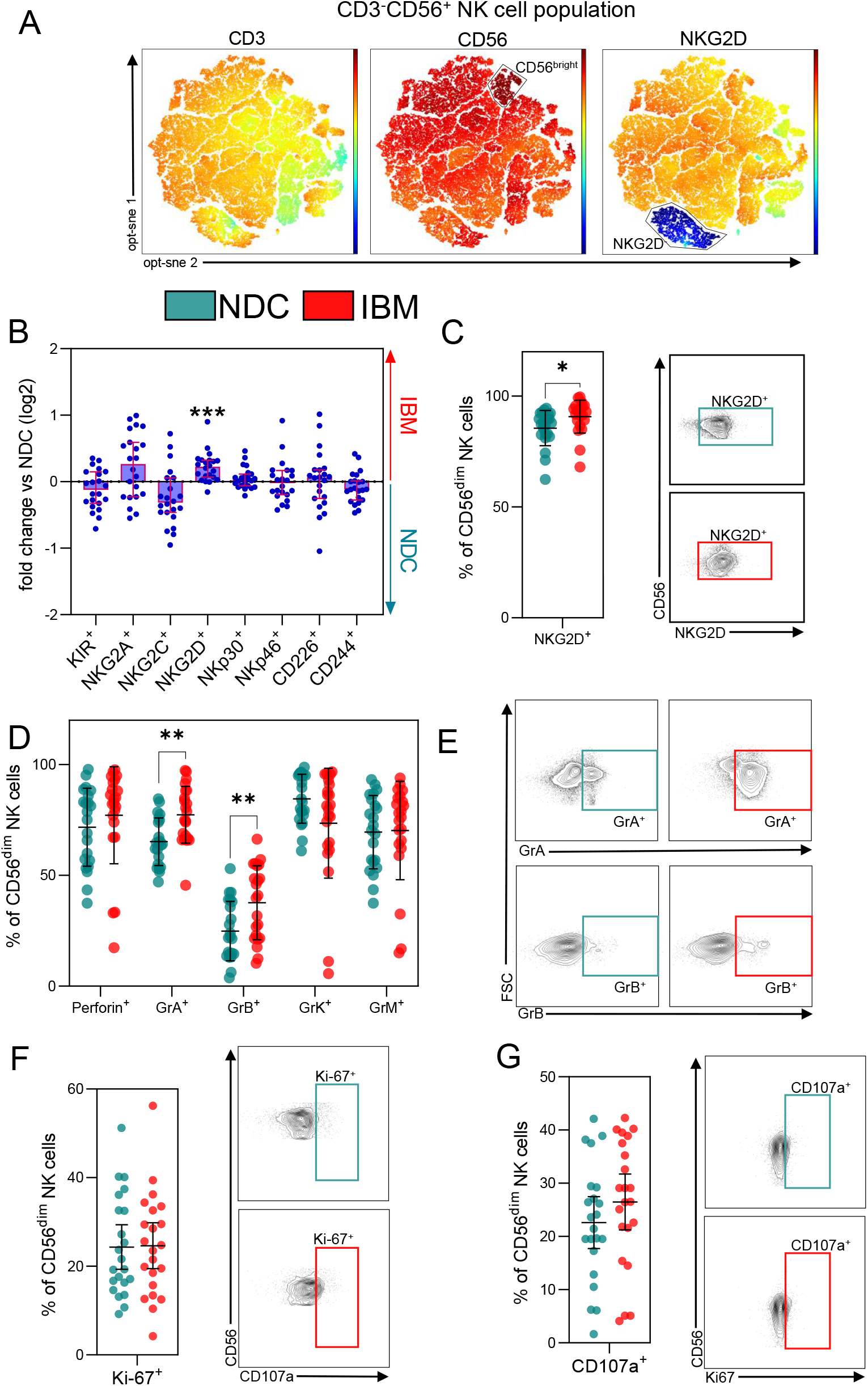
NKG2D is upregulated in IBM. We analysed the NK cell compartment as defined by CD^-^CD56^+^ cells. (**A**) Dimensional reduction with opt-sne indicating levels of CD3, CD56 and NKG2D for the NK cell gate. (**B**) Fold change of the MFI comparing IBM to NDC for the indicated NK cell receptors for CD56^dim^ NK cells. Each dot is one patient. Statistical analysis was performed using Kruskal-Wallis one-way analysis. (**C**) Relative cell numbers of NKG2D positive CD56^dim^ NK cells and exemplary gates for the comparison. (**D**) Comparison of the relative numbers of CD56^dim^ NK cells positive for the indicated granzymes or perforins. (**E**) Exemplary gates for GrA and GrB. (**F**) Comparison of the relative numbers of CD56^dim^ NK cells positive for Ki-67 and for (**G**) CD107a with the respective exemplary gates. Differences were determined by the two-sided student’s T-test. Abbreviations: Gr = granzyme, IBM = inclusion body myositis, MFI = mean fluorescence intensity, NDC = non-diseased control, NK = natural killer, PA = pulmonary affection *** p < 0.001, ** p < 0.01

We utilized a multicolor flow cytometry panel consisting of eight surface molecules associated with NK cell activation and cytotoxic potential. Surface levels were quantified by mean fluorescence intensity (MFI). Broadly, NK cells from IBM patients and NDCs showed similar patterns across most markers in this panel (**Fig. 2B**). However, NKG2D was significantly elevated on CD56^dim^ NK cells from IBM patients compared to NDC. This was evident both in terms of MFI and the proportion of NKG2D^+^ cells (**Fig. 2C**). As an activating receptor, NKG2D can trigger NK cell cytotoxicity, suggesting a primed cytotoxic state in IBM NK cells ^10,11^.

To further characterize the cytolytic potential of CD56^dim^ NK cells, we performed intracellular staining for granule-associated serine proteases. Compared to NDCs, CD56^dim^ NK cells from IBM patients exhibited markedly higher frequencies of Granzyme A^+^ and Granzyme B^+^, while levels of other granzymes and perforin were not significantly different (**Fig. 2D, E**). This indicates selective upregulation of cytolytic effectors in IBM NK cells.

We next assessed NK cell proliferation using intracellular staining for Ki-67. The proportion of Ki-67^+^ CD56^dim^ NK cells was comparable between IBM and NDCs, suggesting that proliferation is unchanged (**Fig. 2F**). Finally, we evaluated CD107a surface expression as a marker of degranulation. Although CD107a^+^ CD56^dim^ NK cells were more frequent in IBM, the difference did not reach statistical significance due to inter-individual variability (**Fig. 2G**).

In summary, while peripheral lymphocyte and NK cell frequencies remain stable in IBM, NK cells adopt a cytotoxic phenotype. This is characterized by elevated NKG2D expression, increased granzyme content, and enhanced killing capacity, supporting the notion that NK cells may contribute to muscle fiber injury in IBM through NKG2D-driven cytotoxicity.

## Discussion

In this study, we demonstrate that, although the overall frequency of peripheral NK cells remains unchanged in IBM patients, CD56^dim^ NK cells adopt a distinct cytotoxic phenotype characterized by increased NKG2D levels and elevated granzyme content. These findings extend prior immunophenotyping data and implicate NK cells as active contributors to IBM pathology, potentially through both direct cytotoxicity and promotion of oxidative stress.

Previous deep immunophenotyping in IBM blood reported stable total NK cell frequencies, with no significant expansion or contraction of NK subpopulations in patients versus healthy controls ^4^. Our results corroborate these observations at the bulk level, but also add qualitative changes with the alteration of the activating receptor NKG2D on CD56^dim^ NK cells. CD56^dim^ NK cells are thought to be cytotoxic, while CD56brigt are considered a regulatory counterpart ^9^. While this duality has been challenged by recent data ^3,8^, CD56^dim^ NK cells are known to migrate into inflamed tissues ^12,13^ and their phenotypic activation in the periphery suggests recruitment to affected muscle and a possible role in myofiber injury.

The NKG2D-ligand axis represents a possible mechanism by which NK cells recognize and eliminate stressed or transformed cells. We previously reported that in IIMs, NKG2D ligands, such as MICA/B, were upregulated and described tissue invasion by NKG2D^+^ CD8^+^ T cells ^11^. Following this line of thought, expression of NKG2D ligands on inflamed myofibers might provide a target for NKG2D^+^ NK cells as well creating an environment in which muscle fibers become susceptible to NKG2D-dependent NK cell cytotoxicity. Our finding of increased NKG2D MFI and frequency of NKG2D^+^ CD56^dim^ NK cells in IBM patients aligns with this model and suggests that chronic inflammation in IBM might prime NK cells. In line, intracellular analysis revealed selective upregulation of granzyme A and B – but not other granzymes or perforin – in CD56^dim^ NK cells from IBM patients. Granzymes are central mediators of NK cell-induced apoptosis and can also trigger secondary reactive oxygen species (ROS) accumulation within target cells, amplifying injury. Moreover, NK cells themselves can generate ROS via the NADPH oxidase complex (NOX2) during respiratory burst–like activities, a process well-documented in phagocytes and recently recognized in NK cells ^14^. Thus, heightened granzyme content and NKG2D-driven activation might promote two parallel injurious pathways: direct induction of myofiber apoptosis and ROS-mediated oxidative damage. Indeed, oxidative stress is a hallmark of IBM muscle pathology. Studies report mitochondrial DNA deletions, impaired electron transport chain activity, and accumulation of protein aggregates within myofibers, all of which exacerbate ROS production and disrupt cellular homeostasis ^15^. Our data suggest that NK cells may contribute as a potential source of ROS in IBM, acting synergistically with T cell–mediated mechanisms to maintain a pro-oxidative milieu that impairs muscle regeneration.

Therapeutically, these insights highlight several potential intervention points. Blocking NKG2D signaling – either by inhibiting receptor–ligand interactions or by preventing ligand upregulation on muscle fibers – could attenuate NK cell–mediated cytotoxicity. Similarly, targeted suppression of NK cell activation (e.g., via IL-15 pathway inhibitors) or selective scavenging of ROS in the muscle microenvironment may protect myofibers from oxidative injury.

Several limitations warrant consideration. Our study focused on peripheral blood NK cells; direct analysis of muscle-infiltrating NK cells would verify recruitment and a local activation state. The cross-sectional design precludes assessment of temporal dynamics – longitudinal profiling could reveal whether NK cell activation correlates with disease progression or response to therapy. Finally, the mechanisms by which NK-derived ROS interact with myocellular antioxidant defenses remain to be elucidated and may uncover novel redox-sensitive targets.

In conclusion, our findings suggest that NK cells might amplify muscle fiber damage via NKG2D-driven cytotoxicity and ROS generation. Additional research into their role in the inflamed muscle is needed to provide further data on this concept.

## Acknowledgements

We thank the patients and their families for their support.

## References

1. Lundberg, I. E., de Visser, M. & Werth, V. P. Classification of myositis. Nat Rev Rheumatol 14, 269–278 (2018).

2. Greenberg, S. A. Inclusion body myositis: clinical features and pathogenesis. Nat Rev Rheumatol 15, 257–272 (2019).

3. Wolf, N. K., Kissiov, D. U. & Raulet, D. H. Roles of natural killer cells in immunity to cancer, and applications to immunotherapy. Nat Rev Immunol 23, 90–105 (2023).

4. Goyal, N. A. et al. Immunophenotyping of Inclusion Body Myositis Blood T and NK Cells. Neurology 98, e1374–e1383 (2022).

5. Eschborn, M. et al. Evaluation of Age-Dependent Immune Signatures in Patients With Multiple Sclerosis. Neurol Neuroimmunol Neuroinflamm 8, e1094 (2021).

6. Nelke, C. et al. K2P2.1 is a regulator of inflammatory cell responses in idiopathic inflammatory myopathies. J Autoimmun 142, 103136 (2023).

7. Lilleker, J. B. et al. 272nd ENMC international workshop: 10 Years of progress - revision of the ENMC 2013 diagnostic criteria for inclusion body myositis and clinical trial readiness. 16-18 June 2023, Hoofddorp, The Netherlands. Neuromuscul Disord 37, 36–51 (2024).

8. Weber, S. et al. Distinctive CD56dim NK subset profiles and increased NKG2D expression in blood NK cells of Parkinson’s disease patients. NPJ Parkinsons Dis 10, 36 (2024).

9. De Maria, A., Bozzano, F., Cantoni, C. & Moretta, L. Revisiting human natural killer cell subset function revealed cytolytic CD56(dim)CD16+ NK cells as rapid producers of abundant IFN-gamma on activation. Proc Natl Acad Sci U S A 108, 728–732 (2011).

10. David, C. et al. Impact of NKG2D Signaling on Natural Killer and T-Cell Function in Cerebral Ischemia. J Am Heart Assoc 12, e029529 (2023).

11. Ruck, T. et al. The NKG2D-IL-15 signaling pathway contributes to T-cell mediated pathology in inflammatory myopathies. Oncotarget 6, 43230–43243 (2015).

12. Petri, R. M. et al. Activated Tissue-Resident Mesenchymal Stromal Cells Regulate Natural Killer Cell Immune and Tissue-Regenerative Function. Stem Cell Reports 9, 985–998 (2017).

13. Fu, B., Tian, Z. & Wei, H. Subsets of human natural killer cells and their regulatory effects. Immunology 141, 483–489 (2014).

14. Aydin, E., Johansson, J., Nazir, F. H., Hellstrand, K. & Martner, A. Role of NOX2-Derived Reactive Oxygen Species in NK Cell-Mediated Control of Murine Melanoma Metastasis. Cancer Immunol Res 5, 804–811 (2017).

15. Kleefeld, F. et al. Mitochondrial damage is associated with an early immune response in inclusion body myositis. Brain awaf118 (2025) doi:10.1093/brain/awaf118.

